# Reducing Functional Dysconnectivity in Schizophrenia Spectrum Disorders

**DOI:** 10.1101/2024.09.26.24314430

**Authors:** Stephan Wunderlich, Daniel Keeser, Johanna Spaeth, Isabel Maurus, Cagatay Alici, Andrea Schmitt, Peter Falkai, Sophia Stoecklein, Lukas Roell

## Abstract

**Background:** As a condition of dysconnectivity, schizophrenia spectrum disorders (SSD) are characterized by positive, negative, and cognitive symptoms. To improve these symptoms in SSD, physical exercise interventions show promise. We examined if reductions of functional dysconnectivity following exercise therapy are associated with clinical improvements in SSD and explored potential genetic underpinnings.

**Methods:** The study utilized data from the ESPRIT C3 trial, investigating the effects of aerobic exercise versus flexibility, strengthening, and balance training on different health outcomes in individuals with SSD. Functional dysconnectivity in 23 patients relative to a healthy reference sample, was assessed both pre- and post-intervention. Changes of functional dysconnectivity after exercise and their clinical relevance were evaluated. An imaging transcriptomics approach was used to study the link between changes in functional dysconnectivity and gene expression profiles.

**Results:** We observed substantial reductions of functional dysconnectivity on the whole-brain level linked to enhanced gene expression mainly in oligodendrocytes. With regard to the clinical implications, decreases of dysconnectivity in the default-mode network were associated with improvements in global functioning. Reductions of dysconnectivity within the salience network were linked to improvements in symptom severity. Lastly, reductions of functional dysconnectivity in language regions such as Broca’s area were related to cognitive benefits.

**Conclusions:** Our study supports a recent theory of oligodendrocyte pathology in SSD and suggests that reducing functional dysconnectivity in the default-mode, salience, and language network reflect a potential therapeutic target to improve global functioning, total symptom severity, and cognitive impairments in post-acute SSD.

Trial name: ESPRIT C3

Registry: International Clinical Trials Database, ClinicalTrials.gov Registration number: NCT03466112

URL: https://clinicaltrials.gov/ct2/show/NCT03466112?term=NCT03466112&draw=2&rank=1

## INTRODUCTION

The dysconnection hypothesis of schizophrenia claims that psychosis is characterized by impaired synaptic efficacy throughout the brain, leading to widespread macroscale deteriorations in intrinsic and extrinsic connectivity related to schizophrenia-typical positive, negative, and cognitive symptoms (1). Accordingly, several large-scale neuroimaging studies have demonstrated multiple aberrant functional connectivity patterns in people with schizophrenia spectrum disorders (SSD) compared to healthy controls (2–10). In particular, functional dysconnectivity within and between core brain networks, such as the default-mode, salience, fronto-parietal, somatormotor, limbic, visual, subcortical or dorsal attention network, reflects a stable transdiagnostic neural marker in psychiatry as assessed by functional magnetic resonance imaging (fMRI) (3,4,8,9). Concerning SSD-specific alterations, several particular dysconnectivity patterns have been proposed: For instance, specific seeds within the default-mode, salience, fronto-parietal and limbic networks show functional dysconnectivity in SSD, but not in other psychiatric conditions (3). Moreover, certain functional disturbances within numerous essential circuits, such as the cortico-striato-pallido-thalamo-cortical circuit (3,11–19), have been frequently reported in SSD. Another central epicenter of continuous neural decline throughout the disease course of psychosis is the hippocampal formation (20), whose connections with the prefrontal cortex are reported to be disturbed in SSD (21). Recently, striatal dysconnectivity has become one of the most promising fMRI-based biomarkers of SSD (6,7,22) that even has the potential to predict improvements in positive symptom severity after antipsychotic treatment (6). Accordingly, as part of the cortico-striato-pallido-thalamo-cortical loop, functional hyperconnectivity between the thalamus/basal ganglia and the auditory-somatomotor network contributes to positive symptoms, whereas hypoconnectivity between the thalamus/basal ganglia and the middle frontal gyrus is linked to cognitive deficits (14). Furthermore, negative symptom severity is associated with disturbed functional connectivity within the default-mode network (10). At the same time, deficits in cognitive domains such as processing speed and working memory are linked to functional dysconnectivity within the salience, auditory, somatomotor and visual networks (2). Importantly, cognitive deterioration is strongly related to impairments in social and occupational functioning (23,24) and thus contributes to low recovery rates in SSD (25). Hence, grounded in the dysconnection hypothesis of schizophrenia, the current state of research demonstrates that SSD is characterized by multiple dysconnectivity patterns across the whole brain that are closely intertwined with the main clinical symptoms of psychotic disorders, namely positive and negative symptoms, as well as cognitive deficits accompanied by impairments in daily life functioning.

To achieve further clinical improvements beyond treatment-as-usual, physical exercise interventions have been proposed as an add-on therapy: Large-scale evidence reveals beneficial effects of different types of physical exercise treatments in people with SSD on positive symptoms (26–35), negative symptoms (26–38), cognitive impairments (35,39–41) and daily life functioning (27,29,42). These effects are expected to be driven by multiple structural and functional adaptations of the brain (43), but the underlying neural mechanisms are not fully understood. Preliminary evidence indicates that higher aerobic fitness, as an indicator of regular involvement in physical exercise, is associated with increased functional connectivity within the default-mode network in SSD (44). Concurrently, aerobic exercise may induce functional adaptations in the cortico-striato-pallido-thalamo-cortical loop and the default-mode network (45). However, these studies examined functional connectivity patterns within specific networks relevant to SSD without putting these patterns in relation to a healthy norm. Hence, previous evidence does not allow conclusions to be drawn about potential normalization processes of functional connectivity following physical exercise treatment in SSD. To address this gap, techniques of normative modeling are required. Concerning functional brain connectivity assessed by resting-state fMRI, Stoecklein et al. (46–48) have proposed the dysconnectivity index (DCI) that quantifies the deviation of functional connectivity from a healthy reference norm for each voxel in each subject and that summarizes these deviation scores as a whole-brain measure of functional dysconnectivity. Importantly, the DCI can also be computed for specific brain regions and networks, referred to as the specific DCI.

Aiming to provide further insights in the genetic underpinnings of macroscale neural alterations such as functional dysconnectivity, an imaging transcriptomics approach has been proposed recently (49–51). In this context, such technique enables to explore associations between functional dysconnectivity patterns in patients with SSD and general gene expression profiles in the human brain derived from the Allen Human Brain Atlas (52). Applying imaging transcriptomics has the potential to provide insights into the genetic origins of functional dysconnectivity in SSD, thereby offering a more sophisticated mechanistic understanding of the disorder (49–51).

Combining the DCI with an imaging transcriptomics approach, we investigate connectome-based mechanisms that drive clinical improvements in schizophrenia and study the genetic basis of changes in functional dysconnectivity. A better understanding of such mechanisms may help to develop targeted therapies and guide future treatment decisions.

## METHODS AND MATERIALS

The current work is based on data from the Enhancing Schizophrenia Prevention and Recovery through Innovative Treatments (ESPRIT) C3 study. The ESPRIT C3 study is an already completed multicenter randomized-controlled trial that explored the effects of aerobic exercise compared to flexibility, strengthening and balance training on several health outcomes in people with SSD. Details on the study and the main clinical results are published elsewhere (35,53). The study is in line with the Declaration of Helsinki and ethical approval was provided by the local ethics committee of the Ludwig-Maximilians-University Hospital. With respect to the project at hand, only data that were acquired at the Department of Psychiatry and Psychotherapy and the Department of Radiology of the Ludwig-Maximilians-University Hospital in Munich were utilized.

### Study Design and Sample

For the main ESPRIT C3 study, the intention-to-treat sample consisted of 180 individuals with SSD. Participants were randomly assigned either to aerobic endurance training (AET) or to flexibility, strengthening and balance training (FSBT). Patients in both groups exercised up to three times per week, between 40 and 50 minutes, for six months. In the AET group, participants cycled on a stationary bicycle ergometer at an individualized moderate exercise intensity determined via a stepwise lactate threshold test. Patients in the FSBT group performed various stretching, mobility, stability, balance, and relaxation exercises. Details regarding the study design, randomization process, blinding procedures, organization of the exercise training, and other relevant methodological considerations are described elsewhere (35,53).

Regarding the present work, only data from subjects who underwent structural and functional magnetic resonance imaging (MRI) before and after six months of the intervention were considered. After quality control, a total of 23 people with SSD were considered in the final statistical analyses. The healthy reference sample comprised 200 individuals (120 female, 80 male), with an average age of 30.20 years (± 3.19 years), using data from the Genomics Superstruct Project (GSP) (54).

### MRI Data Acquisition

SSD patients were scanned with a 3T Siemens Magnetom Skyra MRI scanner (SIEMENS Healthineers AG, Erlangen, Germany) at the Department of Radiology of the Ludwig-Maximilians-University Hospital Munich. One 3D T1-weighted magnetization-prepared rapid gradient echo (MPRAGE) sequence with an isotropic spatial resolution of 0.8 x 0.8 x 0.8 mm^3^ and two resting-state functional echo planar imaging (EPI) sequences with a total duration of twelve minutes were acquired. Supplementary Table S1 displays the scanning parameters. The scanning protocol utilized for the GSP data closely resembles that of the current scanning protocol.

### Quality Control and Preprocessing

Structural and resting-state fMRI data were quality controlled based on both visual inspection of images before and after preprocessing and the automated quality control software MRIQC (55). All data achieved after preprocessing had a temporal signal-to-noise ratio (tSNR) above 100 and a mean framewise displacement (mFD) below 0.3 mm.

Structural MRI data were processed using FreeSurfer version 6.0 (http://surfer.nmr.mgh.harvard.edu). Preprocessing of the resting-state fMRI was performed using fMRIPrep version 20.2.2 (56). The first five functional frames were removed, while the remaining frames were standardized and smoothed with a 6.0 mm Full Width at Half Maximum Gaussian kernel. MCFLIRT (FSL v5.0.9) was applied for the subsequent head motion correction using ICA-AROMA (57). Lastly, nuisance regression, filtering and detrending were performed.

For the whole-brain DCI, blood-oxygenation level-dependent signals were extracted for each subject, and correlation matrices were calculated separately for the left hemisphere (3352 voxels) and right hemisphere (3316 voxels), followed by z-standardization. In the case of the calculation of the specific DCI, correlation matrices were computed based on blood-oxygenation level-dependent signals from specific seeds representing the respective brain regions and networks of interest and subsequently z-standardized. Further details on data preprocessing are described in the supplemental information.

### Calculation of the Whole-Brain DCI

The DCI was computed similarly to approaches outlined in prior work by Stoecklein et al. (46), as detailed in the supplemental information. In essence, connections deviating beyond a specific threshold from the distribution observed in the reference group were classified as “dysconnected”. The dysconnectivity count (DCC) for each patient was then aggregated within each hemisphere and normalized by the number of voxels in that hemisphere to yield the whole-brain DCI.

### Calculation of the DCI within Specific Brain Regions and Networks

In order to probe subtle changes in brain connectivity, we extracted pertinent brain voxels from both the reference group and schizophrenia patients. We examined these voxels across multiple networks, encompassing the visual, somatomotor, limbic, fronto-parietal, default-mode, dorsal attention, salience, and subcortical networks. Additionally, we explored connectivity within specific brain regions of interest, such as the hippocampal formation and between the hippocampus and the prefrontal cortex, the thalamus and the middle frontal gyrus and the thalamus and the somatomotor network. Following that, we exclusively calculated the DCC for each of these designated brain regions and networks, yielding a specific DCI that encapsulates these specific functional dysconnectivity patterns.

### Assessment of Whole-Brain DCC Changes and Clinical Outcomes

The Global Assessment of Functioning (GAF) scale was administered to assess the general status of psychiatric symptoms and functioning (58). To cover total schizophrenia symptom severity, consisting of positive and negative symptoms, as well as general psychopathology, the Positive and Negative Syndrome Scale (PANSS) (59) was utilized. Aiming to measure global cognitive performance, several cognitive tests that target different subdomains of cognition (e.g. short- and long-term memory, working memory, verbal fluency, sustained attention) were performed and summarized to a composite score (for details see supplemental information). All clinical measures were acquired at baseline before the start of the intervention and post intervention after six months of physical exercise. The change scores between baseline and post-intervention were calculated in such a way that positive values represent an improvement and negative values represent a deterioration on the respective scale. The whole-brain DCC of each voxel was normalized across all subjects and the change scores were computed by subtracting the DCC for each voxel at baseline from the respective DCC at follow-up. Hence, a positive DCC change score reflected a reduction of dysconnectivity in the corresponding voxel.

### Statistical Data Analysis

Aiming to examine if whole-brain and specific functional dysconnectivity in people with SSD decreases after physical exercise treatment, we calculated 13 linear mixed effect models for repeated measures using the *lme4* package (60) in R v4.2.2. Session (baseline, six months), age, sex, chlorpromazine equivalents, number of achieved exercise sessions, group (AET, FSBT) and hemisphere (left, right) were included as fixed effects, while the whole-brain and specific DCI served as dependent variables. A random intercept was included for each subject. The p-values of the predictor *Session* were extracted and corrected for multiple testing across the 13 linear mixed effect models using the False Discovery Rate (FDR) method. In the case of significance (q < 0.05) after FDR correction, Tukey post-hoc tests for the particular DCI outcome were calculated to identify the direction and size of the effect, expressed as Coheńs d and the respective 95% confidence interval.

To study associations between clinical changes in the GAF, PANSS, and cognitive composite scores on the one hand and changes in the whole-brain DCC of each voxel on the other hand, we performed voxel-wise Pearson’s correlations. Following this, we projected the correlation values onto a normative brain. This approach enabled us to investigate how changes in clinical measures aligned with dysconnectivity patterns in the brain across the aforementioned assessment scales.

### Imaging Transcriptomics Approach

We first created DCC brain maps in the MNI standard space at 2mm isotropic resolution, indicating the severity of dysconnectivity for each subject in each voxel at both timepoints. Using the *glm* module in Nilearn version 0.10.4, we computed a voxel-wise ANOVA for repeated measures including session (baseline, six months), age, sex, chlorpromazine equivalents, number of achieved exercise sessions and group (AET, FSBT) as factors. We derived continuous statistical brain maps indicating the voxel-wise change of dysconnectivity from pre- to post-intervention across the whole sample. These maps were parcellated according to the Brainnetome atlas (61) using the *parcellate* module from neuromaps (62). Analogously, we parcellated the Allen Human Brain Atlas based on the Brainnetome atlas in MNI space at an isotropic resolution of 2mm using the abagen toolbox (63). Based on the resampled and parcellated statistical DCC brain map, we generated respective null models according to Burt et al. (2020) (64) to preserve spatial autocorrelation and computed Spearman correlations between DCC changes and gene expression data utilizing the *stats* module from neuromaps (62). This resulted in a vector of correlation coefficients for each gene included in the Allen Human Brain Atlas, revealing the correlation between functional dysconnectivity changes in patients and the gene-specific expression in the human brain across all regions defined in the

Brainnetome atlas. To gain an insight in the types of genes that reveal consistent associations with changes in functional dysconnectivity, we used this vector to calculate a gene set enrichment analysis based on the corresponding gene-to-cell-type annotations provided by Lake et al. (2018) (65) using the gseapy toolbox (66).

## RESULTS

### Patients Characteristics

The majority of the final study sample was male, middle-aged, well-educated, and in the post-acute phase of the disease. Moreover, most of the participants considered here were randomized to the FSBT exercise group, having on average completed 1.3 exercise sessions per week. The detailed characteristics of the study sample are summarized in Table 1.

**Table 1.** Sample Characteristics.

| | **Patients with SSD**<br>**N = 23**<br>(n/mean ± SD) |
| --- | --- |
| **Exercise group** |  |
| AET | 9 (39.1 %) |
| FSBT | 14 (60.9 %) |
| **Sex** |  |
| Female | 7 (30.4 %) |
| Male | 16 (69.6 %) |
| **Age (years)** | 37.17 ± 10.38 |
| **PANSS at baseline** |  |
| Positive symptoms | 11.17 ± 3.39 |
| Negative symptoms | 10.65 ± 3.31 |
| Total symptoms | 45.74 ± 9.48 |
| **GAF at baseline** | 64.57 ± 7.79 |
| **Chlorpromazine equivalents** | 428.04 ± 243.76 |
| **Education (years)** | 16.30 ± 4.16 |
| **Total number of trainings** | 34.52 ± 15.86 |

The sample size refers to the number of participants that were considered in the final statistical data analysis. Chlorpromazine equivalents were computed according to the Defined Daily Doses method. N, total sample size; n, sample size per category; AET, aerobic endurance training; FSBT, flexibility, strengthening and balance training; PANSS, Positive and Negative Syndrome Scale; GAF, Global Assessment of Functioning Scale; SD, standard deviation.

### Changes of Whole-Brain and Specific Functional Dysconnectivity after Physical Exercise

The predictor *Time* was significant after FDR correction for the following whole-brain and specific DCI variants: whole brain (q < 0.001), visual network (q < 0.001), somatomotor network (q < 0.001), limbic network (q < 0.001), default-mode network (q < 0.001), salience network (q < 0.001), subcortical network (q < 0.001), thalamic-prefrontal connection (q < 0.001), thalamic-somatomotor connection (q < 0.001), hippocampal-prefrontal connection (q < 0.001), and connectivity within the hippocampal formation (q < 0.001). These effects indicated changes over time in the corresponding DCI values across both exercise groups. There was no significant change in the specific DCI for the fronto-parietal and dorsal attention network. Tukey post-hoc tests demonstrated significant reductions from baseline to post-intervention of the whole-brain DCI (d = −2.73, CI = [−3.15, −2.31], p_tukey_ < 0.001) and the specific DCI of the somatomotor network (d = −2.52, CI = [−2.94, −2.12], p_tukey_ < 0.001), limbic network (d = −1.87, CI = [−2.28, − 1.45], p_tukey_ < 0.001), default-mode network (d = −2.19, CI = [−2.61, −1.78], p_tukey_ < 0.001), salience network (d = −1.84, CI = [−2.25, −1.42], p_tukey_ < 0.001), subcortical network (d = −1.18, CI = [−1.59, −0.76], p_tukey_ < 0.001), thalamic-middle frontal connection (d = −1.47, CI = [−1.89, −1.05], p_tukey_ < 0.001), thalamic-somatomotor connection (d = −2.25, CI = [−2.67, −1.84], p_tukey_ < 0.001), hippocampal-prefrontal connection (d = −1.34, CI = [−1.76, −0.92], p_tukey_ < 0.001), and connectivity within the hippocampal formation (d = −2.19, CI = [−2.61, −1.78], p_tukey_ < 0.001). Increases in the DCI were observed for the visual network (d = 1.45, CI = [1.03, 1.86], p_tukey_ < 0.001). Figure 1 illustrates the time effects on all computed DCI scores. The full test statistics of the other fixed effects are summarized in Table S2.

**Figure 1.**
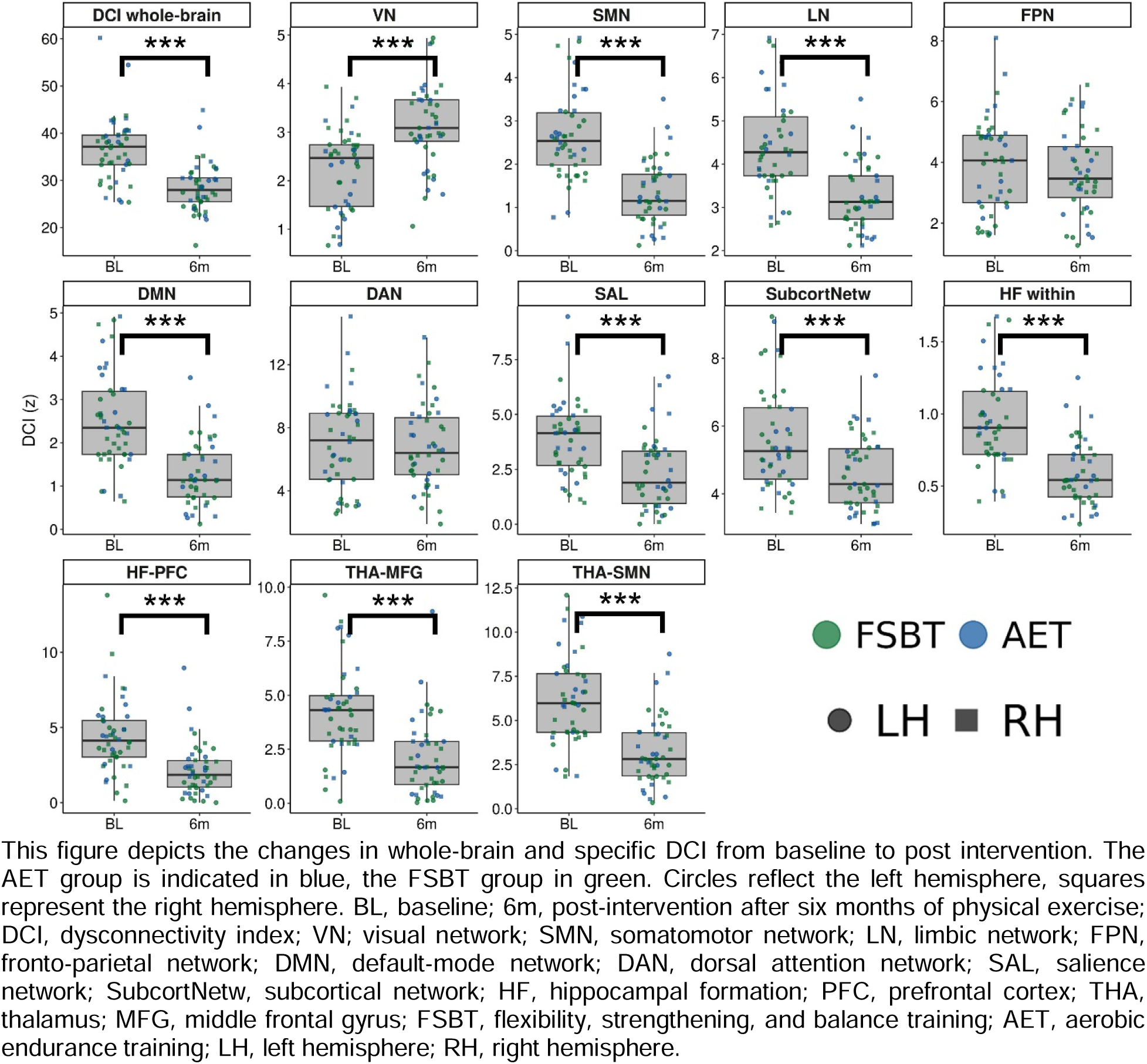
Changes in Whole-Brain and Specific DCI.

### Brain Region and Network Specific Correlations Between Reductions of Whole-Brain DCC and Clinical Improvements

Figure 2A depicts the strongest correlation between improvement in the GAF and adaptations in the DCC in the default-mode network on a voxel-wise level. Dysconnectivity reduction in insular regions is associated with improvements in GAF and PANSS. Furthermore, dysconnectivity in somatomotor and frontoparietal networks predominantly correlates with improvements in the PANSS score (2B). Additionally, improved cognition predominantly manifests through a reduction in DCC, notably observed in left hemisphere language regions such as Broca’s areas (2C).

**Figure 2.**
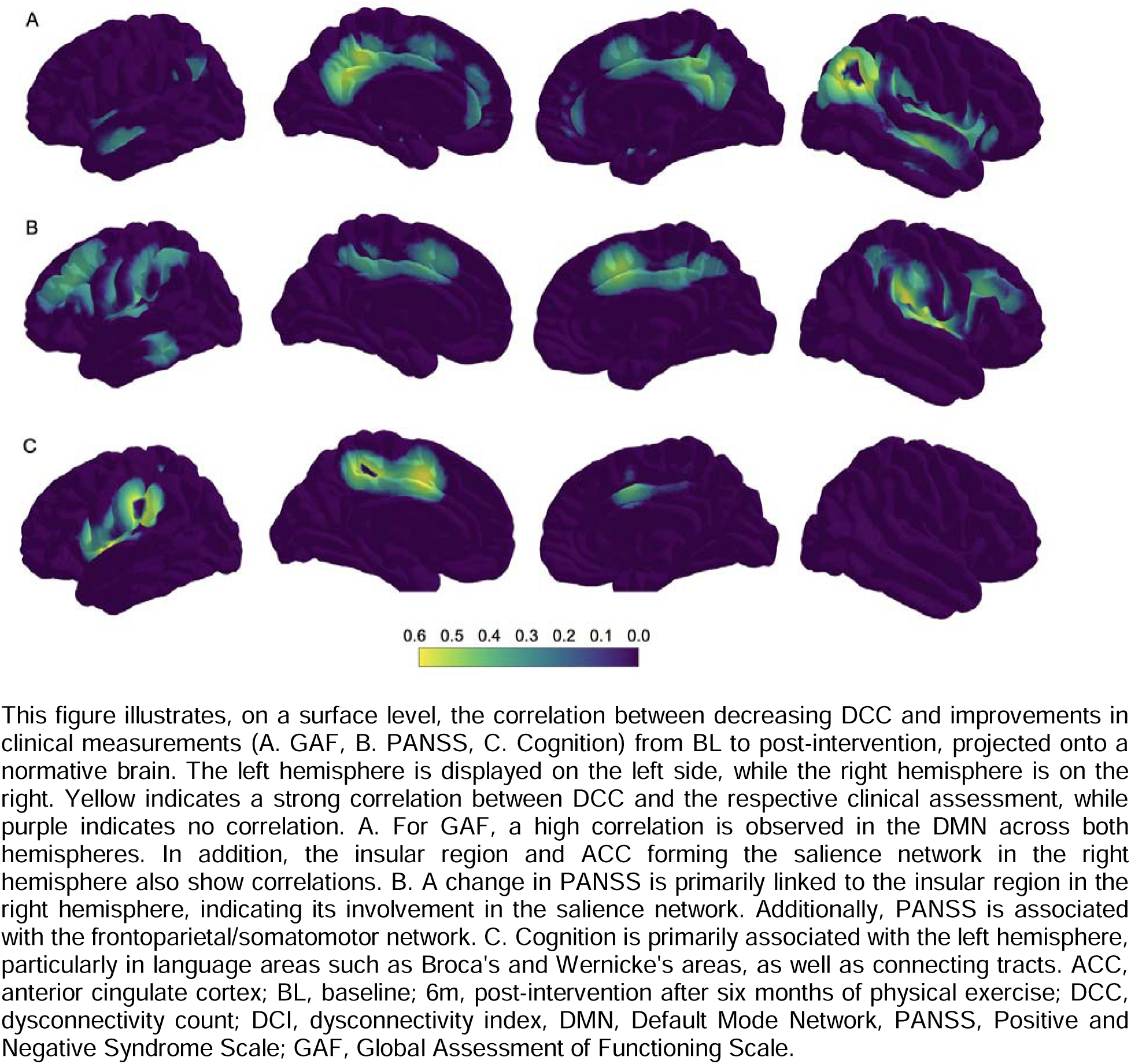
Correlation Between DCC Change and Change in Clinical Measurements.

### Association between Changes of Functional Dysconnectivity and Gene Expression

Figure 3 summarizes the results from the gene set enrichment analysis. Across brain regions, we obtained that reductions of functional dysconnectivity from baseline to post-intervention were linked to higher expression of genes typically enriched in oligodendrocytes, oligodendrocyte precursor cells (OPCs), astrocytes, endothelial cells, pericytes, and microglia. Contrastingly, reductions of functional dysconnectivity from baseline to post-intervention were associated with lower expression of genes typically enriched in excitatory and inhibitory neurons.

**Figure 3.**
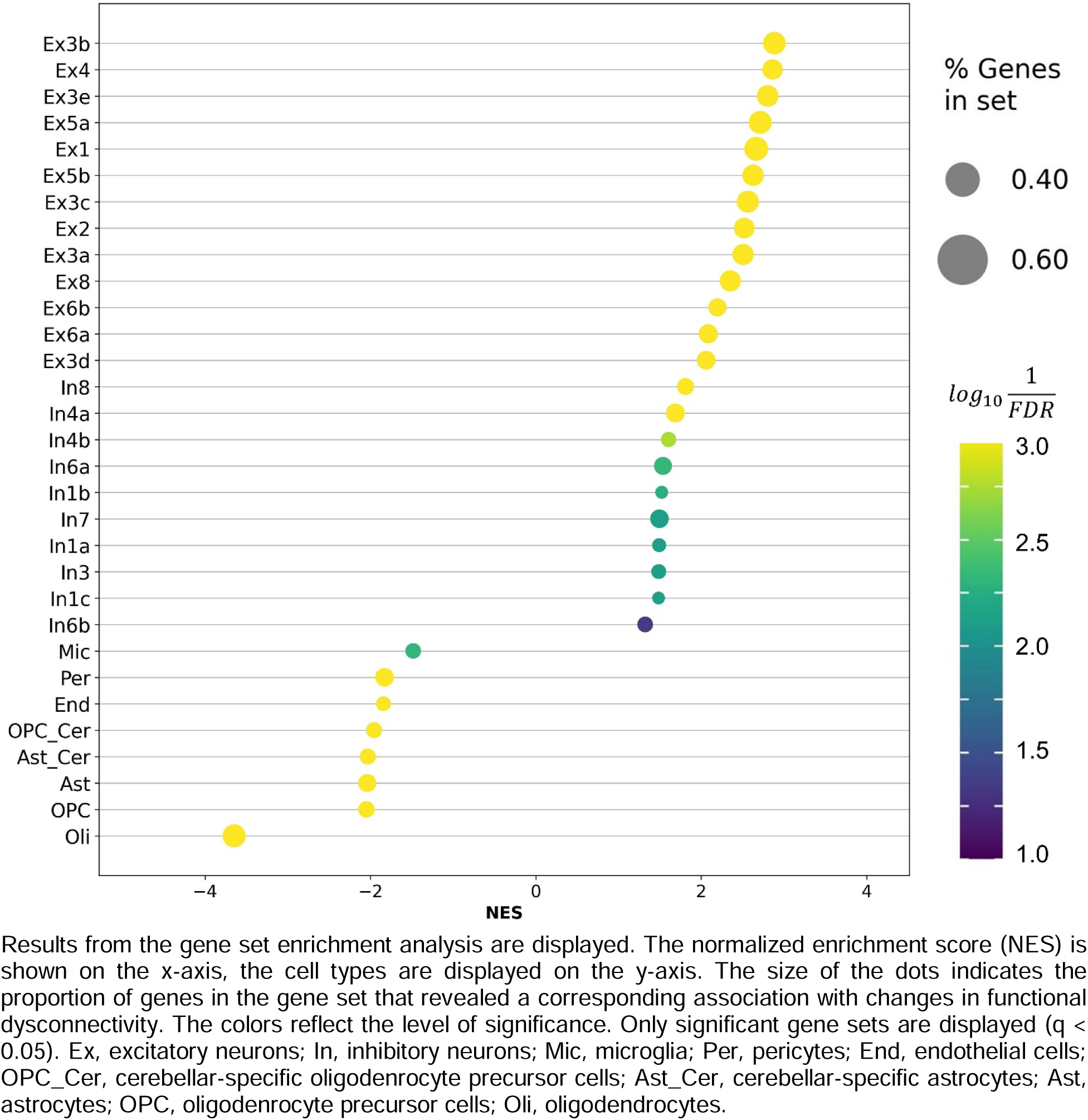
Gene set enrichment analysis.

## DISCUSSION

The current study explored the clinical relevance of potential decreases in functional dysconnectivity after physical exercise treatment in people with SSD and the respective genetic underpinnings, aiming to identify connectome-based mechanisms that drive clinical improvements.

We observed substantial reductions in the whole-brain DCI after physical exercise therapy in SSD. To our knowledge, this is the first time such a finding has been reported. Previous results in manifested SSD and in individuals with clinical high-risk for psychosis suggest adaptations of single functional connections induced by aerobic exercise interventions within the default-mode network, the cortico-striato-pallido-thalamo-cortical loop or between hippocampal and occipital regions (45,67,68). These studies are in line with our current results, as they indicate that functional connectivity patterns in SSD can generally change in the context of a physical exercise intervention. However, direct comparisons to the current findings remain challenging due to substantial differences in methodology. Particularly, the distinctive approach of the DCI in contrast to conventional measures of functional connectivity provides unique insights into individual-specific deviations of functional connectivity. In our case, reductions of functional dysconnectivity after physical exercise can be interpreted as normalization processes of the individual brains of patients. Such inference is not possible when only focusing on functional connectivity changes, as performed in previous studies (45,67,68). From a clinical standpoint, compared to previous research, our objective was to investigate individual alterations in functional dysconnectivity over time and under two distinct exercise interventions. However, previous studies have primarily compared a moderate aerobic exercise intervention either against another form of exercise regimen (45,67) or against a waitlist control (68). Hence, these examinations aimed to investigate potential adaptations in functional connectivity particularly induced by aerobic exercise interventions compared to a control condition. In contrast, we were interested in potential changes in functional dysconnectivity after physical exercise treatment without claiming that the substantial reductions we obtained were specifically induced by exercise. Consequently, the large decrease in functional dysconnectivity observed in our data may result from a complex interaction of several factors the patients experienced in the course of their study participation, such as social, psychological, exercise or even placebo effects. Thus, based on our data, the current substantial reductions in whole-brain functional dysconnectivity are not attributable solely to the effects of physical exercise but reveal that such global interventions can be used to counteract functional brain dysconnectivity in SSD. This is of particular importance because SSD is seen as a disorder of dysconnectivity both on the theoretical (1) and on the empirical level (3).

Beyond reductions in whole-brain functional dysconnectivity, we also found specific decreases in multiple networks and regions relevant to SSD, such as the default-mode network, salience network, somatomotor network, limbic network, subcortical network, prefrontal-thalamic-somatomotor pathway, hippocampal-prefrontal connections, and within the hippocampal formation. Aiming to cover the clinical implications of changes in functional dysconnectivity, our findings suggest that reductions of functional dysconnectivity in the default-mode network were linked to improvements in global functioning. The default-mode network is generally associated with self-referential and interoceptive behavior, theory of mind, and social cognition (69,70). In the psychiatric context, disorder-related alterations within the default-mode network are linked to negative symptom severity in people with SSD (10) and to impaired inhibition control in several psychiatric disorders (9). Our previous analysis indicated that aerobic exercise-induced volume increases in the posterior cingulate gyrus, a central node of the default-mode network, were linked to an improvement in general disorder severity (45). These results reveal the broad and diverse functionality of the default-mode network in both healthy individuals and psychiatric patients. Our findings on the associations between modifications of dysconnectivity in the default-mode network and changes in global levels of functioning correspond to this notion. In sum, we propose that functional dysconnectivity within the default-mode network may reflect a promising neural target for future therapeutic approaches to improve the global level of functioning in people with SSD. Targeted treatment candidates may be brain stimulation approaches such as focused ultrasound or transcranial magnetic stimulation that both have the potential to modulate functional connectivity (71,72).

Our findings further indicate that a reduction in functional dysconnectivity in insular regions and the somatomotor network was associated with ameliorations in total symptom severity. Alterations of the insula in SSD have been associated with the impaired ability of patients to process representations of themselves, leading to a disturbed discrimination between self-generated and external information (73). As the latter is supposed to contribute to two main symptoms observed in people with SSD, namely hallucinations and delusions, our current findings on the relation between reductions in insular dysconnectivity and improvements in clinical symptoms appear plausible. With respect to the functional connectivity of the somatomotor network, previous evidence also suggests associations with different symptom domains of SSD (11,14,74,75). Hence, we conclude that reducing dysconnectivity in the insula and the somatomotor network through targeted therapeutic interventions such as aforementioned brain stimulation techniques (71,72) may represent a promising strategy to achieve ameliorations in the total symptom severity of patients with SSD.

Lastly, a decrease in functional dysconnectivity in language regions on the left hemisphere such as Broca’s areas was related to cognitive improvements. To our knowledge, this is the first time such a finding has been described in people with SSD. In general, parts of Brocás area are part of a broader left-hemispheric language network involved in language processing, while other parts contribute to wider cognitive functions such as explicit and working memory (69,76). Given that the cognitive composite score used in the current study comprised several cognitive tests that required language and memory functions (see supplemental information), the observed link between dysconnectivity changes in Brocás area and cognitive adaptations seems reasonable. Consequently, achieving decreases in functional dysconnectivity in Brocás area using respective brain stimulation approaches (71,72) may help to increase cognitive benefits in future treatments for people with SSD.

Delving into the genetic underpinnings of reductions of functional dysconnectivity after physical exercise in SSD, we observed that these reductions were most strongly associated with a higher expression of genes typically enriched in oligodendrocytes and OPCs. In other words: In brain regions that show stronger reductions of functional dysconnectivity, the expression of genes related to oligodendrocytes and OPCs is generally more pronounced. This is the first time such finding has been reported. However, a recent theory of the etiology of cognitive deficits in SSD claims that an erroneous maturation of OPCs into oligodendrocytes leads to impaired myelination and disturbed micro-und macroscale connectivity manifesting in global cognitive deteriorations (77). This theory is based on a wide range of different findings supporting the involvement of OPCs and oligodendrocytes in the pathophysiology of schizophrenia: For instance, among other cell types, OPCs and oligodendrocytes were partly enriched in genes linked to SSD (78), while several single genes that play a prominent role for the functioning of OPCs and oligodendrocytes have been related to reduced structural connectivity in SSD (77). In addition, several stereology studies reveal reductions of the number of oligodendrocytes in people with SSD compared to controls (79–81). Our current results can be embedded in this line of evidence, as they indicate that reductions of functional dysconnectivity in SSD mainly occur in brain regions in which the expression of genes related to OPCs and oligodendrocytes is higher. This may point toward a remyelination process that may have occurred in the context of the physical exercise intervention, although such inferences should be interpreted with caution given the limitations of an imaging transcriptomics approach (49–51).

Despite these promising findings, we acknowledge that our study includes several important limitations that provide essential implications for future research. First, only 23 patients with SSD could be included in the final analysis. The study population, which the current analysis is based on, is at a rather stable phase of the disease, preventing the generalizability of our current findings to a broader spectrum of patients with SSD. Especially the current very large effect sizes regarding the dysconnectivity reductions may be rather sample-specific and will probably not be obtained in more representative patient samples. Therefore, we emphasize that our results require replication in an independent and larger sample of people with SSD assessed in a comparable interventional design. Second, due to a lack of a control group of healthy individuals we could not control for technical scanner variance and natural fluctuations of functional dysconnectivity patterns potentially occurring between both measurement timepoints. However, we assume that potential natural fluctuations of functional dysconnectivity that may be present in healthy cohorts cannot explain the substantial and systematic effects obtained in this analysis. Nonetheless, future studies of this type should include a longitudinal cohort of healthy controls, aiming to systematically account for technical variability and natural changes in functional dysconnectivity. Third, the voxel-wise correlations between adaptations in functional dysconnectivity and clinical changes reflect rather descriptive associations without systematic significance testing and correction for multiple comparisons. We decided to follow a more liberal statistical approach as the first step, aiming to generate hypotheses for subsequent studies. Hence, future hypothesis-driven research approaches in this area should evaluate if the present associations between clinical changes and adaptations in functional dysconnectivity in the default-mode network, insula, somatomotor network and Brocás area are reproducible.

To conclude, our study indicates that decreases of functional dysconnectivity within the default-mode network is closely tied to ameliorations in global functioning. Reductions in functional dysconnectivity within the salience and somatomotor network correlate with improvements in total symptom severity. Similarly, a decrease in functional dysconnectivity within Broca’s area corresponds with improved cognitive functioning. Observed reductions of functional dysconnectivity are related to an increased expression of genes enriched in oligodendrocytes and oligodendrocyte precursor cells. Our results provide first insights into potential connectome-based therapeutic targets in post-acute patients with SSD. In particular, future therapeutic approaches could aim to reduce functional dysconnectivity in the default-mode network, insula, somatomotor network, and Brocás area to achieve multifaceted improvements on the clinical level.

## Data Availability

All data produced in the present study are available upon reasonable request to the authors.

## ACKNOWLEDGMENTS

The work was supported by the German Federal Ministry of Education and Research (BMBF) through the research network on psychiatric diseases ESPRIT (Enhancing Schizophrenia Prevention and Recovery through Innovative Treatments; coordinator: Andreas Meyer-Lindenberg; grant number, 01EE1407E) to PF and AS. Furthermore, the study was supported by the Else Kröner-Fresenius Foundation with the Research College “Translational Psychiatry” to PF, AS, and IM (Residency/PhD track of the International Max Planck Research School for Translational Psychiatry [IMPRS-TP]). The study was endorsed by the Federal Ministry of Education and Research (Bundesministerium für Bildung und Forschung [BMBF]) within the initial phase of the German Center for Mental Health (DZPG) (grant: 01EE2303A, 01EE2303F to PF, AS). The authors express their appreciation to the Clinical Open Research Engine (CORE) at the University Hospital LMU (Munich, Germany) for providing the computational infrastructure to run the CPU-intensive MRI analysis pipelines.

## DISCLOSURES

PF is a co-editor of the German (DGPPN) schizophrenia treatment guidelines and a co-author of the WFSBP schizophrenia treatment guidelines; he is on the advisory boards and receives speaker fees from Janssen, Lundbeck, Otsuka, Servier, and Richter. SW, DK, JS, IM, CA, AS, SS and LR declare no conflicts of interest or financial disclosures relevant to this research.

## Notes

### Author Declarations

Ethical approval was provided by the local ethics committee of the Ludwig-Maximilians-University Hospital.

## REFERENCES

1. Friston K, Brown HR, Siemerkus J, Stephan KE (2016): The dysconnection hypothesis (2016). Schizophrenia Research 176: 83–94.

2. Adhikari BM, Hong LE, Sampath H, Chiappelli J, Jahanshad N, Thompson PM, et al. (2019): Functional network connectivity impairments and core cognitive deficits in schizophrenia. Human Brain Mapping 40: 4593–4605.

3. Brandl F, Avram M, Weise B, Shang J, Simões B, Bertram T, et al. (2019): Specific Substantial Dysconnectivity in Schizophrenia: A Transdiagnostic Multimodal Meta-analysis of Resting-State Functional and Structural Magnetic Resonance Imaging Studies. Biological Psychiatry 85: 573– 583.

4. Dong D, Wang Y, Chang X, Luo C, Yao D (2018): Dysfunction of Large-Scale Brain Networks in Schizophrenia: A Meta-analysis of Resting-State Functional Connectivity. Schizophrenia Bulletin 44: 168–181.

5. Li S, Hu N, Zhang W, Tao B, Dai J, Gong Y, et al. (2019): Dysconnectivity of Multiple Brain Networks in Schizophrenia: A Meta-Analysis of Resting-State Functional Connectivity. Front Psychiatry 10: 482.

6. Li A, Zalesky A, Yue W, Howes O, Yan H, Liu Y, et al. (2020): A neuroimaging biomarker for striatal dysfunction in schizophrenia. Nat Med 26: 558–565.

7. Rubio JM, Lencz T, Cao H, Kraguljac N, Dhamala E, Homan P, et al. (2024): Replication of a neuroimaging biomarker for striatal dysfunction in psychosis. Mol Psychiatry. 10.1038/s41380-023-02381-9

8. Goodkind M, Eickhoff SB, Oathes DJ, Jiang Y, Chang A, Jones-Hagata LB, et al. (2015): Identification of a Common Neurobiological Substrate for Mental Illness. JAMA Psychiatry 72: 305.

9. Sha Z, Wager TD, Mechelli A, He Y (2019): Common Dysfunction of Large-Scale Neurocognitive Networks Across Psychiatric Disorders. Biological Psychiatry 85: 379–388.

10. O’Neill A, Mechelli A, Bhattacharyya S (2019): Dysconnectivity of Large-Scale Functional Networks in Early Psychosis: A Meta-analysis. Schizophrenia Bulletin 45: 579–590.

11. Anticevic A, Cole MW, Repovs G, Murray JD, Brumbaugh MS, Winkler AM, et al. (2014): Characterizing thalamo-cortical disturbances in schizophrenia and bipolar illness. Cereb Cortex 24: 3116–3130.

12. Anticevic A, Haut K, Murray JD, Repovs G, Yang GJ, Diehl C, et al. (2015): Association of Thalamic Dysconnectivity and Conversion to Psychosis in Youth and Young Adults at Elevated Clinical Risk. JAMA Psychiatry 72: 882–891.

13. Anticevic A, Yang G, Savic A, Murray JD, Cole MW, Repovs G, et al. (2014): Mediodorsal and visual thalamic connectivity differ in schizophrenia and bipolar disorder with and without psychosis history. Schizophr Bull 40: 1227–1243.

14. Avram M, Brandl F, Bäuml J, Sorg C (2018): Cortico-thalamic hypo- and hyperconnectivity extend consistently to basal ganglia in schizophrenia. Neuropsychopharmacology 43: 2239–2248.

15. Lui S, Yao L, Xiao Y, Keedy SK, Reilly JL, Keefe RS, et al. (2015): Resting-state brain function in schizophrenia and psychotic bipolar probands and their first-degree relatives. Psychol Med 45: 97–108.

16. Tu P-C, Lee Y-C, Chen Y-S, Hsu J-W, Li C-T, Su T-P (2015): Network-specific cortico-thalamic dysconnection in schizophrenia revealed by intrinsic functional connectivity analyses. Schizophr Res 166: 137–143.

17. Welsh RC, Chen AC, Taylor SF (2010): Low-frequency BOLD fluctuations demonstrate altered thalamocortical connectivity in schizophrenia. Schizophr Bull 36: 713–722.

18. Woodward ND, Karbasforoushan H, Heckers S (2012): Thalamocortical dysconnectivity in schizophrenia. Am J Psychiatry 169: 1092–1099.

19. Woodward ND, Heckers S (2016): Mapping Thalamocortical Functional Connectivity in Chronic and Early Stages of Psychotic Disorders. Biol Psychiatry 79: 1016–1025.

20. Chopra S, Segal A, Oldham S, Holmes A, Sabaroedin K, Orchard ER, et al. (2023): Network-Based Spreading of Gray Matter Changes Across Different Stages of Psychosis. JAMA Psychiatry 80: 1246.

21. Sigurdsson T, Duvarci S (2016): Hippocampal-Prefrontal Interactions in Cognition, Behavior and Psychiatric Disease. Front Syst Neurosci 9. 10.3389/fnsys.2015.00190

22. Mueller S, Wang D, Pan R, Holt DJ, Liu H (2015): Abnormalities in hemispheric specialization of caudate nucleus connectivity in schizophrenia. JAMA Psychiatry 72: 552–560.

23. Tsang HWH, Leung AY, Chung RCK, Bell M, Cheung W-M (2010): Review on vocational predictors: a systematic review of predictors of vocational outcomes among individuals with schizophrenia: an update since 1998. Aust N Z J Psychiatry 44: 495–504.

24. Ventura J, Hellemann GS, Thames AD, Koellner V, Nuechterlein KH (2009): Symptoms as mediators of the relationship between neurocognition and functional outcome in schizophrenia: a meta-analysis. Schizophr Res 113: 189–199.

25. Van Eck RM, Burger TJ, Vellinga A, Schirmbeck F, de Haan L (2018): The Relationship Between Clinical and Personal Recovery in Patients With Schizophrenia Spectrum Disorders: A Systematic Review and Meta-analysis. Schizophr Bull 44: 631–642.

26. Firth J, Cotter J, Elliott R, French P, Yung AR (2015): A systematic review and meta-analysis of exercise interventions in schizophrenia patients. Psychol Med 45: 1343–1361.

27. Dauwan M, Begemann MJH, Heringa SM, Sommer IE (2016): Exercise Improves Clinical Symptoms, Quality of Life, Global Functioning, and Depression in Schizophrenia: A Systematic Review and Meta-analysis. SCHBUL 42: 588–599.

28. Wei G-X, Yang L, Imm K, Loprinzi PD, Smith L, Zhang X, Yu Q (2020): Effects of Mind–Body Exercises on Schizophrenia: A Systematic Review With Meta-Analysis. Front Psychiatry 11: 819.

29. Fernández-Abascal B, Suárez-Pinilla P, Cobo-Corrales C, Crespo-Facorro B, Suárez-Pinilla M (2021): In- and outpatient lifestyle interventions on diet and exercise and their effect on physical and psychological health: a systematic review and meta-analysis of randomised controlled trials in patients with schizophrenia spectrum disorders and first episode of psychosis. Neuroscience & Biobehavioral Reviews 125: 535–568.

30. Bredin SSD, Kaufman KL, Chow MI, Lang DJ, Wu N, Kim DD, Warburton DER (2022): Effects of Aerobic, Resistance, and Combined Exercise Training on Psychiatric Symptom Severity and Related Health Measures in Adults Living With Schizophrenia: A Systematic Review and Meta-Analysis. Front Cardiovasc Med 8: 753117.

31. Ziebart C, Bobos P, MacDermid JC, Furtado R, Sobczak DJ, Doering M (2022): The efficacy and safety of exercise and physical activity on psychosis: A systematic review and meta-analysis. Front Psychiatry 13: 807140.

32. Gallardo-Gómez D, Noetel M, Álvarez-Barbosa F, Alfonso-Rosa RM, Ramos-Munell J, Del Pozo Cruz B, Del Pozo-Cruz J (2023): Exercise to treat psychopathology and other clinical outcomes in schizophrenia: A systematic review and meta-analysis. Eur Psychiatr 66: e40.

33. Guo J, Liu K, Liao Y, Qin Y, Yue W (2024): Efficacy and feasibility of aerobic exercise interventions as an adjunctive treatment for patients with schizophrenia: a meta-Analysis. Schizophr 10: 2.

34. Rißmayer M, Kambeitz J, Javelle F, Lichtenstein TK (2024): Systematic Review and Meta-analysis of Exercise Interventions for Psychotic Disorders: The Impact of Exercise Intensity, Mindfulness Components, and Other Moderators on Symptoms, Functioning, and Cardiometabolic Health. Schizophr Bull sbae015.

35. Maurus I, Roell L, Lembeck M, Papazova I, Greska D, Muenz S, et al. (2023): Exercise as an add-on treatment in individuals with schizophrenia: Results from a large multicenter randomized controlled trial. Psychiatry Research 328: 115480.

36. Vogel JS, Van Der Gaag M, Slofstra C, Knegtering H, Bruins J, Castelein S (2019): The effect of mind-body and aerobic exercise on negative symptoms in schizophrenia: A meta-analysis. Psychiatry Research 279: 295–305.

37. Sabe M, Kaiser S, Sentissi O (2020): Physical exercise for negative symptoms of schizophrenia: Systematic review of randomized controlled trials and meta-analysis. General Hospital Psychiatry 62: 13–20.

38. Kim M, Lee Y, Kang H (2023): Effects of Exercise on Positive Symptoms, Negative Symptoms, and Depression in Patients with Schizophrenia: A Systematic Review and Meta-Analysis. IJERPH 20: 3719.

39. Firth J, Stubbs B, Rosenbaum S, Vancampfort D, Malchow B, Schuch F, et al. (2016): Aerobic Exercise Improves Cognitive Functioning in People With Schizophrenia: A Systematic Review and Meta-Analysis. SCHBUL sbw115.

40. Shimada T, Ito S, Makabe A, Yamanushi A, Takenaka A, Kawano K, Kobayashi M (2022): Aerobic exercise and cognitive functioning in schizophrenia: An updated systematic review and meta-analysis. Psychiatry Res 314: 114656.

41. Xu Y, Cai Z, Fang C, Zheng J, Shan J, Yang Y (2022): Impact of aerobic exercise on cognitive function in patients with schizophrenia during daily care: A meta-analysis. Psychiatry Research 312: 114560.

42. Korman N, Stanton R, Vecchio A, Chapman J, Parker S, Martland R, et al. (2023): The effect of exercise on global, social, daily living and occupational functioning in people living with schizophrenia: A systematic review and meta-analysis. Schizophr Res 256: 98–111.

43. Maurus I, Hasan A, Röh A, Takahashi S, Rauchmann B, Keeser D, et al. (2019): Neurobiological effects of aerobic exercise, with a focus on patients with schizophrenia. Eur Arch Psychiatry Clin Neurosci 269: 499–515.

44. Roell L, Maurus I, Keeser D, Karali T, Papazov B, Hasan A, et al. (2022): Association between aerobic fitness and the functional connectome in patients with schizophrenia. Eur Arch Psychiatry Clin Neurosci 272: 1253–1272.

45. Roell L, Keeser D, Papazov B, Lembeck M, Papazova I, Greska D, et al. (n.d.): Effects of Exercise on Structural and Functional Brain Patterns in Schizophrenia— Data From a Multicenter Randomized-Controlled Study.

46. Stoecklein VM, Stoecklein S, Galiè F, Ren J, Schmutzer M, Unterrainer M, et al. (2020): Resting-state fMRI detects alterations in whole brain connectivity related to tumor biology in glioma patients. Neuro Oncol 22: 1388–1398.

47. Stoecklein VM, Wunderlich S, Papazov B, Thon N, Schmutzer M, Schinner R, et al. (2023): Perifocal Edema in Patients with Meningioma is Associated with Impaired Whole-Brain Connectivity as Detected by Resting-State fMRI. AJNR Am J Neuroradiol 44: 814–819.

48. Stoecklein S, Wunderlich S, Papazov B, Winkelmann M, Kunz WG, Mueller K, et al. (2023): Functional connectivity MRI provides an imaging correlate for chimeric antigen receptor T-cell-associated neurotoxicity. Neurooncol Adv 5: vdad135.

49. Martins D, Giacomel A, Williams SCR, Turkheimer F, Dipasquale O, Veronese M, PET Templates Working Group (2021): Imaging transcriptomics: Convergent cellular, transcriptomic, and molecular neuroimaging signatures in the healthy adult human brain. Cell Rep 37: 110173.

50. Fornito A, Arnatkevičiūtė A, Fulcher BD (2019): Bridging the Gap between Connectome and Transcriptome. Trends Cogn Sci 23: 34–50.

51. Arnatkeviciute A, Fulcher BD, Fornito A (2019): A practical guide to linking brain-wide gene expression and neuroimaging data. Neuroimage 189: 353–367.

52. Hawrylycz MJ, Lein ES, Guillozet-Bongaarts AL, Shen EH, Ng L, Miller JA, et al. (2012): An anatomically comprehensive atlas of the adult human brain transcriptome. Nature 489: 391–399.

53. Maurus I, Hasan A, Schmitt A, Roeh A, Keeser D, Malchow B, et al. (2021): Aerobic endurance training to improve cognition and enhance recovery in schizophrenia: design and methodology of a multicenter randomized controlled trial. Eur Arch Psychiatry Clin Neurosci 271: 315–324.

54. Holmes AJ, Hollinshead MO, O’Keefe TM, Petrov VI, Fariello GR, Wald LL, et al. (2015): Brain Genomics Superstruct Project initial data release with structural, functional, and behavioral measures. Sci Data 2: 150031.

55. Esteban O, Birman D, Schaer M, Koyejo OO, Poldrack RA, Gorgolewski KJ (2017): MRIQC: Advancing the automatic prediction of image quality in MRI from unseen sites. PLoS One 12: e0184661.

56. Esteban O, Markiewicz CJ, Blair RW, Moodie CA, Isik AI, Erramuzpe A, et al. (2019): fMRIPrep: a robust preprocessing pipeline for functional MRI. Nat Methods 16: 111–116.

57. Pruim RHR, Mennes M, Buitelaar JK, Beckmann CF (2015): Evaluation of ICA-AROMA and alternative strategies for motion artifact removal in resting state fMRI. Neuroimage 112: 278–287.

58. Endicott J, Spitzer RL, Fleiss JL, Cohen J (1976): The global assessment scale. A procedure for measuring overall severity of psychiatric disturbance. Arch Gen Psychiatry 33: 766–771.

59. Kay SR, Fiszbein A, Opler LA (1987): The positive and negative syndrome scale (PANSS) for schizophrenia. Schizophr Bull 13: 261–276.

60. Bates D, Mächler M, Bolker B, Walker S (2015): Fitting Linear Mixed-Effects Models Using lme4. J Stat Soft 67: 1–48.

61. Fan L, Li H, Zhuo J, Zhang Y, Wang J, Chen L, et al. (2016): The Human Brainnetome Atlas: A New Brain Atlas Based on Connectional Architecture. Cereb Cortex 26: 3508–3526.

62. Markello RD, Hansen JY, Liu Z-Q, Bazinet V, Shafiei G, Suárez LE, et al. (2022): neuromaps: structural and functional interpretation of brain maps. Nat Methods 19: 1472–1479.

63. Markello RD, Arnatkeviciute A, Poline J-B, Fulcher BD, Fornito A, Misic B (2021): Standardizing workflows in imaging transcriptomics with the abagen toolbox. Elife 10: e72129.

64. Burt JB, Helmer M, Shinn M, Anticevic A, Murray JD (2020): Generative modeling of brain maps with spatial autocorrelation. Neuroimage 220: 117038.

65. Lake BB, Chen S, Sos BC, Fan J, Kaeser GE, Yung YC, et al. (2018): Integrative single-cell analysis of transcriptional and epigenetic states in the human adult brain. Nat Biotechnol 36: 70–80.

66. Fang Z, Liu X, Peltz G (2023): GSEApy: a comprehensive package for performing gene set enrichment analysis in Python. Bioinformatics 39: btac757.

67. Dean DJ, Bryan AD, Newberry R, Gupta T, Carol E, Mittal VA (2017): A Supervised Exercise Intervention for Youth at Risk for Psychosis: An Open-Label Pilot Study. J Clin Psychiatry 78: e1167–e1173.

68. Damme KSF, Gupta T, Ristanovic I, Kimhy D, Bryan AD, Mittal VA (2022): Exercise Intervention in Individuals at Clinical High Risk for Psychosis: Benefits to Fitness, Symptoms, Hippocampal Volumes, and Functional Connectivity. Schizophrenia Bulletin 48: 1394–1405.

69. Laird AR, Fox PM, Eickhoff SB, Turner JA, Ray KL, McKay DR, et al. (2011): Behavioral interpretations of intrinsic connectivity networks. J Cogn Neurosci 23: 4022–4037.

70. Mak LE, Minuzzi L, MacQueen G, Hall G, Kennedy SH, Milev R (2017): The Default Mode Network in Healthy Individuals: A Systematic Review and Meta-Analysis. Brain Connect 7: 25–33.

71. Chang K-Y, Tik M, Mizutani-Tiebel Y, Schuler A-L, Taylor P, Campana M, et al. (2024): Neural response during prefrontal theta burst stimulation: Interleaved TMS-fMRI of full iTBS protocols. Neuroimage 291: 120596.

72. Yaakub SN, White TA, Roberts J, Martin E, Verhagen L, Stagg CJ, et al. (2023): Transcranial focused ultrasound-mediated neurochemical and functional connectivity changes in deep cortical regions in humans. Nat Commun 14: 5318.

73. Wylie KP, Tregellas JR (2010): The role of the insula in schizophrenia. Schizophr Res 123: 93–104.

74. Cheng W, Palaniyappan L, Li M, Kendrick KM, Zhang J, Luo Q, et al. (2015): Voxel-based, brain-wide association study of aberrant functional connectivity in schizophrenia implicates thalamocortical circuitry. NPJ Schizophr 1: 15016.

75. Li T, Wang Q, Zhang J, Rolls ET, Yang W, Palaniyappan L, et al. (2016): Brain-Wide Analysis of Functional Connectivity in First-Episode and Chronic Stages of Schizophrenia. SCHBUL sbw099.

76. Fedorenko E, Blank IA (2020): Broca’s Area Is Not a Natural Kind. Trends Cogn Sci 24: 270–284.

77. Falkai P, Rossner MJ, Raabe FJ, Wagner E, Keeser D, Maurus I, et al. (2023): Disturbed Oligodendroglial Maturation Causes Cognitive Dysfunction in Schizophrenia: A New Hypothesis. Schizophr Bull 49: 1614–1624.

78. Skene NG, Bryois J, Bakken TE, Breen G, Crowley JJ, Gaspar HA, et al. (2018): Genetic identification of brain cell types underlying schizophrenia. Nat Genet 50: 825–833.

79. Schmitt A, Steyskal C, Bernstein H-G, Schneider-Axmann T, Parlapani E, Schaeffer EL, et al. (2009): Stereologic investigation of the posterior part of the hippocampus in schizophrenia. Acta Neuropathol 117: 395–407.

80. Falkai P, Malchow B, Wetzestein K, Nowastowski V, Bernstein H-G, Steiner J, et al. (2016): Decreased Oligodendrocyte and Neuron Number in Anterior Hippocampal Areas and the Entire Hippocampus in Schizophrenia: A Stereological Postmortem Study. Schizophr Bull 42 Suppl 1: S4–S12.

81. Schmitt A, Tatsch L, Vollhardt A, Schneider-Axmann T, Raabe FJ, Roell L, et al. (2022): Decreased Oligodendrocyte Number in Hippocampal Subfield CA4 in Schizophrenia: A Replication Study. Cells 11: 3242.

