## Supplemental information for "Reducing Functional Dysconnectivity in Schizophrenia Spectrum Disorders"

**Supplementary Information**

**Cognitive Test Batteries**

We administered the Verbal Learning and Memory Test (VLMT) (1), Digit Span Test (DST) (2), Trail Making Test (TMT) A and B (3), the category naming part of the Brief Cognitive Assessment Tool for Schizophrenia (B-CATS) (4), the Digit Symbol Substitution Test (DSST) (2), and an adjusted version of the Emotion Recognition Test (ERT) (5).

Within the VLMT the investigator reads a list of 15 words and the participants had to remember as many words as possible in arbitrary order. This procedure was repeated five times in a row (VLMT-1^st^ to VLMT-5^th^) and the sum of correctly remembered words across the five trials was computed (VLMT-sum). After the fifth trial, an interference list of 15 different words was read and the subjects had to name as many words from this new list as possible (VLMT-inter). The subject was asked to remember as many words as possible from the first list (VLMT-6^th^) without repeating it again. After a 20 minutes delay in which other cognitive tests were executed, the participants had to remember as many words as possible from the first list again (VLMT-7^th^). The difference in remembered words between VLMT-5^th^ and VLMT-7^th^ was calculated and multiplied by minus one (VLMT-diff). Finally, the investigator reads 50 words including the ones from the first list and the interference trial and the subjects had to decide if the corresponding word was part of the first list (VLMT-recog).

The number of correctly remembered or recognized words in each trial was counted and z-standardized resulting in eight different VLMT-scores. Z-scores from the VLMT-1^st^, VLMT-inter, VLMT-6^th^, VLMT-7^th^ and VLMT-diff were averaged to a mean VLMT score.

During the DST-forward the investigator read digit rows of increasing lengths that the subject had to repeat verbally in the same order. The test was stopped if the participant failed twice within the same “row length category”. DST-backward worked analogously, except that the subjects had to repeat the digits in reverse order. The number of correct trials was counted, z-standardized in both versions separately and averaged to a global DST score.

During the TMT-A subjects had to connect numbers from 1 to 25 in ascending order as fast and accurately as possible without lifting the pencil from the sheet of paper. In TMT-B participants had to connect numbers and letters alternately in the following order until number 13 was reached: 1-A-2-B-3-C-4-D-5-E-6-F-7-G-8-H-9-I-10-J-11-K-12-L-13. The time needed in seconds was measured. Results from both versions were multiplied by minus one, z-standardized across sessions and participants and averaged to a global TMT score.

The task in the category naming part of the B-CATS was to name as many animals (B-CATS-animals), fruits (B-CATS-fruits) and vegetables (B-CATS-vegetables) as possible within one minute for each category. The number of correctly named animals, fruits and vegetables was z-standardized and averaged to a global B-CATS score.

In the DSST the subjects were asked to translate as many numbers as possible within 90 seconds into written symbols based on a predefined coding scheme. The number of correctly translated numbers was assessed and z-standardized.

During the ERT emotional faces illustrating anger, fear, happiness, disgust, surprise, sadness, or neutrality were presented to the subjects who had to recognize the correct emotion. The number of correctly recognized emotions was counted and z-standardized.

The final scores of the VLMT, DST, TMT, B-CATS, DSST and ERT were z-standardized across subjects and sessions and averaged to the global cognition score used in the current study.

**Processing of Functional Magnetic Resonance Imaging (FMRI) Data**

First, the orientation of the MRI image was changed to the radiologically conventional LAS (Left-Anterior-Superior) system using fslswapdim, and if necessary, the orientation information of the MRI image was additionally modified using fslorient (Functional Magnetic Resonance Imaging of the Brain, FMRIB, FMRIB Software Library, FSL v5.0.9 (6).

The anatomical segmentation and cortical reconstruction were performed using the software package FreeSurfer Version 6.0 (recon-all command, (<http://surfer.nmr.mgh.harvard.edu/>). FMRI images were preprocessed using fMRIPrep version 20.2.2 (7). The preprocessed anatomical data were used utilizing the recon-all command previously executed in FreeSurfer. Head motion correction was performed with ICA-AROMA, including automatic determination of independent components (8). The BOLD signal was filtered using temporal bandpass filters (high-pass = 0.01 Hz, low-pass = 0.08 Hz). Subsequently, potential trends in the data, which may arise from instabilities in the scanner's magnetic field, were removed. After this processing, all data met the criteria for quality control of tSNR > 100 and FD < 0.3 mm.

**Calculation of the Whole-Brain Dysconnectivity Index (DCI) and the Specific DCI**

The calculation of whole-brain DCI was performed on the blood-oxygenation level-dependent (BOLD) signals for the whole left and right hemispheres both at the cortical and subcortical volume level (9). The calculation of the specific DCI is only based on the BOLD signals in pertinent brain voxels being part of specific brain regions or networks of interest. The respective BOLD signals of each patient were compared to the BOLD signals of a healthy cohort of 200 subjects (120 female, 80 male, aged 30.20 years ± 3.19 years), referencing data from the Genomics Superstruct Project (GSP) (10). This dataset includes fMRI and structural MRI data from healthy young adults aged 18 to 35. To closely match the age distribution of our SSD sample, we selected only the oldest participants from the GSP data. This dataset has been utilized in various studies as a healthy reference cohort (11–13). Participants were excluded if their self-reported health information indicated a current or past history of Axis I pathology or neurological disorders, current psychotropic medication usage, acute physical illness, or atypical brain anatomy.

(1) Dysconnectivity Score (DCS)

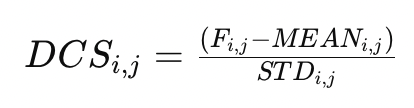

Each i, j where i, j are different voxels. Here F represents the correlation value of the BOLD signal of each voxel i to j of the individual patient. MEAN and STD represent the arithmetic mean and standard deviation of the correlation values between the BOLD signal of voxel i and j of all healthy individuals in the GSP dataset. The set of dysconnective connections is defined as the

(2) Dysconnectivity Count (DCC):

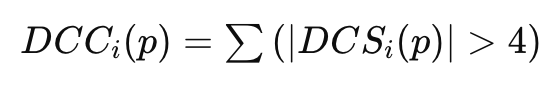

Finally, the quantification of dysconnectivity in each hemisphere, network or specific brain region was performed by summing the dysconnective connections of each voxel, normalized by the number of voxels in each hemisphere, specific brain region or network (3):

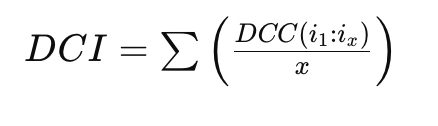

Resulting in a dimensionless whole-brain DCI or a specific DCI per network or of a specific brain region.

**Scanning parameters**

**Table S1***.* Scanning parameters.

| **Site** | **Sequence** | **Resolution** | **TR** | **TE** | **TI** | **FA** | **Slices** | **Timepoints** |
| --- | --- | --- | --- | --- | --- | --- | --- | --- |
| Munich | MP-RAGE | 0.8 × 0.8 × 0.8 mm³ | 2060 ms | 2.2 ms | 1040 ms | 12° | 256 | 248 |
| Munich | EPI | 3.0 × 3.0 × 3.0 mm³ | 3000 ms | 30 ms | - | 85° | 45 | 124 |

Site, study site; Sequence, type of scanning sequence; FoV, field of view; resolution, voxel size; TR, Time of repetition; TE, echo time; TI, inversion time; FA, flip angle; slices, number of acquired slices; MP-RAGE, T1-weighted magnetization prepared rapid gradient echo; EPI, echo planar imaging.

**Formula and full test statistics from linear mixed effect models**

The linear mixed effect model can be expressed as:

$${DCI}_{ijk} = \beta_{0} + \beta_{1}*{Session}_{ijk} + \beta_{2}*{Group}_{i} + \beta_{3}*{Age}_{i} + \beta_{4}*{Sex}_{i} + \beta_{5}*{CPZ}_{i}+ \beta_{6}*{Trainings}_{i} + \beta_{7}*{Education}_{i} + \beta_{8}*{Hemisphere}_{ijk}+ \gamma_{i}+ \epsilon_{ijk}$$

Where ${DCI}_{i,j,k}$ represents the observed DCI of the i^th^ subject at the j^th^ time point in the k^th^ hemisphere. $\beta_{0}$ reflects the intercept of the whole model, while the $\beta_{n}*X_{ijk}$ terms represent the values of fixed effects with their coefficients for the i^th^ subject (i = 1, …, n) at the j^th^ time point (j = 1, 2) in the k^th^ hemisphere (k = 1, 2). $\gamma_{i}$ captures the random intercept of the i^th^ subject. $\epsilon_{ijk}$ is the residual error term.

**Table S2***.* Test statistics.

| **DCI outcome** | **Fixed effect** | **F** | **p_raw_** | **p_raw_ < 0.05** |
| --- | --- | --- | --- | --- |
| total | session | 171.43 | 0.000 | TRUE |
| total | group | 0.88 | 0.363 | FALSE |
| total | age | 0.04 | 0.849 | FALSE |
| total | sex | 0.08 | 0.784 | FALSE |
| total | cpz | 1.94 | 0.182 | FALSE |
| total | education_years | 2.31 | 0.148 | FALSE |
| total | train_num | 0.07 | 0.789 | FALSE |
| total | hemisphere | 12.65 | 0.001 | TRUE |
| VN | session | 48.26 | 0.000 | TRUE |
| VN | group | 0.78 | 0.390 | FALSE |
| VN | age | 0.13 | 0.723 | FALSE |
| VN | sex | 0.01 | 0.943 | FALSE |
| VN | cpz | 0.42 | 0.527 | FALSE |
| VN | education_years | 0.19 | 0.671 | FALSE |
| VN | train_num | 0.09 | 0.767 | FALSE |
| VN | hemisphere | 22.01 | 0.000 | TRUE |
| SMN | session | 146.40 | 0.000 | TRUE |
| SMN | group | 1.95 | 0.181 | FALSE |
| SMN | age | 0.02 | 0.883 | FALSE |
| SMN | sex | 1.42 | 0.250 | FALSE |
| SMN | cpz | 0.68 | 0.421 | FALSE |
| SMN | education_years | 0.51 | 0.487 | FALSE |
| SMN | train_num | 2.07 | 0.169 | FALSE |
| SMN | hemisphere | 0.68 | 0.414 | FALSE |
| LN | session | 80.25 | 0.000 | TRUE |
| LN | group | 2.59 | 0.127 | FALSE |
| LN | age | 0.07 | 0.794 | FALSE |
| LN | sex | 0.48 | 0.497 | FALSE |
| LN | cpz | 1.55 | 0.231 | FALSE |
| LN | education_years | 1.94 | 0.183 | FALSE |
| LN | train_num | 1.75 | 0.205 | FALSE |
| LN | hemisphere | 3.33 | 0.073 | FALSE |
| FPN | session | 0.80 | 0.374 | FALSE |
| FPN | group | 0.29 | 0.596 | FALSE |
| FPN | age | 0.30 | 0.593 | FALSE |
| FPN | sex | 1.77 | 0.202 | FALSE |
| FPN | cpz | 0.00 | 0.945 | FALSE |
| FPN | education_years | 0.48 | 0.497 | FALSE |
| FPN | train_num | 3.78 | 0.070 | FALSE |
| FPN | hemisphere | 24.29 | 0.000 | TRUE |
| DMN | session | 110.43 | 0.000 | TRUE |
| DMN | group | 2.22 | 0.156 | FALSE |
| DMN | age | 0.04 | 0.847 | FALSE |
| DMN | sex | 0.73 | 0.405 | FALSE |
| DMN | cpz | 1.24 | 0.281 | FALSE |
| DMN | education_years | 1.64 | 0.218 | FALSE |
| DMN | train_num | 2.28 | 0.150 | FALSE |
| DMN | hemisphere | 0.59 | 0.447 | FALSE |
| DAN | session | 0.32 | 0.571 | FALSE |
| DAN | group | 0.22 | 0.644 | FALSE |
| DAN | age | 0.97 | 0.340 | FALSE |
| DAN | sex | 0.91 | 0.354 | FALSE |
| DAN | cpz | 0.01 | 0.907 | FALSE |
| DAN | education_years | 0.04 | 0.837 | FALSE |
| DAN | train_num | 1.20 | 0.291 | FALSE |
| DAN | hemisphere | 28.69 | 0.000 | TRUE |
| SAL | session | 77.53 | 0.000 | TRUE |
| SAL | group | 2.51 | 0.133 | FALSE |
| SAL | age | 0.10 | 0.759 | FALSE |
| SAL | sex | 0.45 | 0.514 | FALSE |
| SAL | cpz | 5.56 | 0.031 | TRUE |
| SAL | education_years | 5.98 | 0.026 | TRUE |
| SAL | train_num | 3.34 | 0.086 | FALSE |
| SAL | hemisphere | 2.66 | 0.108 | FALSE |
| SubcortNetw | session | 31.88 | 0.000 | TRUE |
| SubcortNetw | group | 0.23 | 0.635 | FALSE |
| SubcortNetw | age | 0.00 | 0.962 | FALSE |
| SubcortNetw | sex | 0.32 | 0.578 | FALSE |
| SubcortNetw | cpz | 6.48 | 0.022 | TRUE |
| SubcortNetw | education_years | 17.74 | 0.001 | TRUE |
| SubcortNetw | train_num | 5.38 | 0.034 | TRUE |
| SubcortNetw | hemisphere | 6.14 | 0.016 | TRUE |
| THA-MFG | session | 49.77 | 0.000 | TRUE |
| THA-MFG | group | 2.89 | 0.108 | FALSE |
| THA-MFG | age | 0.00 | 0.971 | FALSE |
| THA-MFG | sex | 0.36 | 0.559 | FALSE |
| THA-MFG | cpz | 0.33 | 0.573 | FALSE |
| THA-MFG | education_years | 1.21 | 0.288 | FALSE |
| THA-MFG | train_num | 2.25 | 0.153 | FALSE |
| THA-MFG | hemisphere | 1.20 | 0.277 | FALSE |
| THA-SMN | session | 116.65 | 0.000 | TRUE |
| THA-SMN | group | 2.09 | 0.167 | FALSE |
| THA-SMN | age | 0.10 | 0.761 | FALSE |
| THA-SMN | sex | 0.64 | 0.435 | FALSE |
| THA-SMN | cpz | 1.69 | 0.212 | FALSE |
| THA-SMN | education_years | 1.93 | 0.184 | FALSE |
| THA-SMN | train_num | 2.65 | 0.123 | FALSE |
| THA-SMN | hemisphere | 0.88 | 0.352 | FALSE |
| HF within | session | 110.43 | 0.000 | TRUE |
| HF within | group | 2.22 | 0.156 | FALSE |
| HF within | age | 0.04 | 0.847 | FALSE |
| HF within | sex | 0.73 | 0.405 | FALSE |
| HF within | cpz | 1.24 | 0.281 | FALSE |
| HF within | education_years | 1.64 | 0.218 | FALSE |
| HF within | train_num | 2.28 | 0.150 | FALSE |
| HF within | hemisphere | 0.59 | 0.447 | FALSE |
| HF-PFC | session | 41.26 | 0.000 | TRUE |
| HF-PFC | group | 1.92 | 0.185 | FALSE |
| HF-PFC | age | 0.06 | 0.812 | FALSE |
| HF-PFC | sex | 0.27 | 0.612 | FALSE |
| HF-PFC | cpz | 0.19 | 0.670 | FALSE |
| HF-PFC | education_years | 0.30 | 0.594 | FALSE |
| HF-PFC | train_num | 1.16 | 0.297 | FALSE |
| HF-PFC | hemisphere | 0.14 | 0.711 | FALSE |

DCI, dysconnectivity index; VN; visual network; SMN, somatomotor network; LN, limbic network; FPN, fronto-parietal network; DMN, default-mode network; DAN, dorsal attention network; SAL, salience network; SubcortNetw, subcortical network; HF, hippocampal formation; PFC, prefrontal cortex; THA, thalamus; MFG, middle frontal gyrus.

**Supplemental References**

1. Helmstaedter C, Durwen HF (1990): The Verbal Learning and Retention Test. A useful and differentiated tool in evaluating verbal memory performance. *Schweiz Arch Neurol Psychiatr (1985)* 141: 21–30.

2. Tewes U (1994): *Hamburg-Wechsler Intelligenztest Für Erwachsene Revision 1991 (HAWIE-R)*, vol. 2. Bern: Huber. Retrieved from https://books.google.de/books?id=s4kMYAAACAAJ

3. Reitan R, Wolfson D (1985): *The Halstead-Reitan Neuropsychological Test Battery: Theory and Clinical Interpretation.* Tucson: Neuropsychology Press.

4. Hurford IM, Marder SR, Keefe RS, Reise SP, Bilder RM (2011): A brief cognitive assessment tool for schizophrenia: construction of a tool for clinicians, 2009/09/25 ed. *Schizophrenia Bulletin* 37: 538–545.

5. Ekman P, Friesen WV (1974): Detecting deception from the body or face. *J Pers Soc Psychol* 29: 288–298.

6. Jenkinson M, Bannister P, Brady M, Smith S (2002): Improved optimization for the robust and accurate linear registration and motion correction of brain images. *Neuroimage* 17: 825–841.

7. Esteban O, Markiewicz CJ, Blair RW, Moodie CA, Isik AI, Erramuzpe A, *et al.* (2019): fMRIPrep: a robust preprocessing pipeline for functional MRI. *Nat Methods* 16: 111–116.

8. Pruim RHR, Mennes M, Buitelaar JK, Beckmann CF (2015): Evaluation of ICA-AROMA and alternative strategies for motion artifact removal in resting state fMRI. *Neuroimage* 112: 278–287.

9. Stoecklein VM, Stoecklein S, Galiè F, Ren J, Schmutzer M, Unterrainer M, *et al.* (2020): Resting-state fMRI detects alterations in whole brain connectivity related to tumor biology in glioma patients. *Neuro Oncol* 22: 1388–1398.

10. Holmes AJ, Hollinshead MO, O’Keefe TM, Petrov VI, Fariello GR, Wald LL, *et al.* (2015): Brain Genomics Superstruct Project initial data release with structural, functional, and behavioral measures. *Sci Data* 2: 150031.

11. Yeo BTT, Krienen FM, Sepulcre J, Sabuncu MR, Lashkari D, Hollinshead M, *et al.* (2011): The organization of the human cerebral cortex estimated by intrinsic functional connectivity. *J Neurophysiol* 106: 1125–1165.

12. Buckner RL, Krienen FM, Castellanos A, Diaz JC, Yeo BTT (2011): The organization of the human cerebellum estimated by intrinsic functional connectivity. *J Neurophysiol* 106: 2322–2345.

13. Van Dijk KRA, Sabuncu MR, Buckner RL (2012): The influence of head motion on intrinsic functional connectivity MRI. *Neuroimage* 59: 431–438.
